# Validation of HER2 status in whole genome sequencing data of breast cancers with AI-driven, ploidy-corrected approach

**DOI:** 10.1101/2021.08.30.21258379

**Authors:** Wojtaszewska Marzena, Stępień Rafał, Woźna Alicja, Piernik Maciej, Dąbrowski Maciej, Gniot Michał, Szymański Sławomir, Socha Maciej, Kasprzak Piotr, Matkowski Rafał, Zawadzki Paweł

**Author notes:** **Corresponding author:** Marzena Wojtaszewska, PhD, Central Clinical Hospital of the Internal Affairs in Warsaw, Woloska 137 Str, 02-507 Warsaw, Poland.

## Abstract

The HER2 protein overexpression is one of the most significant biomarkers for breast cancer diagnostics, prediction, and prognostics. The availability of HER2-inhibitors in routine clinical practice directly translates into the diagnostic need for precise and robust marker identification.

At the brink of the genomic era, multigene next-generation sequencing methodologies slowly take over the field of single-biomarker molecular and cytogenetic tests. However, copy number alterations such as amplification of the HER2-coding *ERBB2* gene, are certainly harder to validate as an NGS biomarker than simple SNV mutations. They are characterized by several compound genomic factors i.a. structural heterogeneity, dependence on chromosome count and genomic context of ploidy. In our study, we tested the approach of using whole genome sequencing instead of NGS panels to robustly and accurately determine HER2 status in clinical setup. Based on the large dataset of 877 breast cancer patients’ genomes with curated clinical data and a machine learning approach for optimization of an unbiased diagnostic classifier, we provide a reliable algorithm of HER2 status assessment.

## 1. Introduction

Human epidermal growth factor receptor 2 (HER2) is an important biomarker for targeted therapy in breast cancer (BC). Patients with overexpression of the receptor were considered the worst prognosis group before HER2 inhibitors were introduced into clinical practice [1]. Nowadays, the first and second generation of these drugs slow down disease progression, improving the outcomes in HER2-positive subgroup of BCs. Therefore, it is crucial to accurately and precisely pinpoint the HER2-overexpression status [2].

The molecular mechanism of HER2 overexpression is, in most cases, amplification of a 17q12 chromosome region containing the HER2 coding *ERBB2* gene. The reference method for the assessment of *ERBB2* amplification is immunohistochemistry (IHC) coupled with fluorescence in situ hybridization (FISH) [3]. Currently, diagnostic companies and medical services are beginning to offer novel NGS assays, detecting dozens of actionable biomarkers in a single test. They are trying to incorporate the *ERBB2* copy number (*ERBB2* CN) into their portfolio as well. Unfortunately, *ERBB2* amplification status cannot be easily determined by establishing a simple threshold for negative and positive values, as the genomic context of chromosome 17 copy number and tumour ploidy are interrelated with *ERBB2* CN. Firstly, duplication or triplication of the whole chromosome set or just a subset of chromosomes is a common feature of BC. However, changes in ploidy are seldom associated with overexpression of *ERBB2* gene, as average global transcript levels remain unchanged. Secondly, the isolated deletion or duplication events of chromosome 17 may influence the *ERBB2* transcription [4,5]. The gain of an additional copy of chromosome 17, called polysomy, is correlated with tumour ploidy and is considered its surrogate in the FISH test, but discrepancies between these parameters are in part the reason for inaccuracy in *ERBB2* amplification detection [6].

As it is not feasible to determine ploidy in conventional FISH, the ratio between *ERBB2* CN and chromosome 17 centromeric probe (CEP17) CN serves as a diagnostic criterion in dual probe assays, recommended by official ASCO/CAP Clinical Practice Guidelines for diagnostics of HER2 in breast cancer patients [3].

Whole genome sequencing (WGS) on the other hand is capable of acquiring absolute *ERBB2* copy number, centromere 17 CN and mean ploidy of tumour cells simultaneously. Moreover, WGS can estimate the tumour content of the sample, providing quality control of the material. As WGS is based on PCR-free methodology, it preserves the original proportions of DNA fragments, in contrast to enrichment or PCR-based NGS panels, which may distort the original proportions of DNA fragments and skew the quantification [7].

The purpose of this study was to determine the feasibility of accurately distinguishing between HER2-positive and HER2-negative cases of BC based on matched tumour-normal WGS. Up to date, there have been only a few studies evaluating the clinical utility of NGS testing of *ERBB2* gene status, including WGS method [8–12]. Some of them directly address the clinical need to verify the relevance of their findings for patient management, reporting the overall concordance between IHC/FISH and NGS at about 90% level.

Our study operates on the large population-based cohort of 877 BCs from publicly available databases, supplied with the final clinical HER2 status based on ASCO/CAP guidelines and targeted treatment information, which serves to validate metastatic samples status. We analyzed the whole cohort of patients, aiming to establish the criteria for WGS *ERBB2* status assessment as close to the golden standard as possible, optimized for both sensitivity and precision with a bias-free machine learning approach. We also provide proof-of-concept that genomic data acquired on different platforms with different chemistry yield sufficiently uniform results for molecular diagnostics of ERBB2 amplification by WGS.

## 2. Materials and methods

### 2.1 Sample choice

Matched tumor-normal genomes from 877 breast cancer patients sequenced within three large Genomic Consortia (119-International Cancer Genome Consortium, 70-The Cancer Genome Atlas, 688-Hartwig Medical Foundation; HMF) were downloaded from controlled-access databases after meeting formal criteria [10,13–15]. The samples were sequenced using a low PCR amplification or PCR-free library preparation protocols and paired-end 100-150 base pair Illumina reads with 350-550 base pair insert size (for details, see the Supplementary Table 1). For analyses of primary tumour samples, we included the datasets with clinical HER2 status described as positive or negative, according to ASCO/CAP guidelines 2007-2018 (depending on the year of the original study was conducted, see the Supplementary Table 1). For metastatic/advanced tumour samples from HMF database, metadata on HER2 status were available only for primary tumours, the IHC/FISH status for sequenced sample from second biopsy was not provided. Because of the high rate of conversion from HER2-negative to HER2-positive status (and vice versa) during the cancer evolution [8,9], in metastatic cancers we have taken into consideration also the patients’ treatment metadata and discarded all samples, for which treatment history (pre- and post-biopsy) was discordant with initial HER2 status (eg. if trastuzumab was included in any line of treatment even though HER2 status was reported negative). For details on discarded samples see Supplementary Data.

As there were no new tissue/DNA/RNA samples processed, the written consent of each subject is in possession of data providers. The primary data were collected in accordance with the standards set by the Declaration of Helsinki and the highest data security standards of ISO 27001. There was no need of acquiring approval from a local ethics committee as no actual tumour samples were used.

### 2.2 Whole-genome data processing

The samples were analyzed using publicly available, open-source software embedded within an in-house pipeline (Figure 1) implemented using Ruffus [16]. The analysis started with FASTQ files extraction from the downloaded BAM/CRAM files using Broad Institutes’ Picard [17]. Tumour samples with coverage exceeding 75x were downsampled with Seqtk v1.3-r106 [18] to approx. 60x mean coverage. Next, all reads were trimmed using cutadapt v2.10 [19] and mapped to the GRCh37 genome using Sanger’s Cancerit CGPMAP pipeline v3.0.0 [20]. Samples with uniquely-mapped read coverage below 20x for either tumor or normal genomes were excluded from the analysis [21,22]. Mean tumour samples’ coverage across all datasets after downsampling was 48x, reference blood/EBV-transformed lymphocyte samples’ mean coverage was 36x (detailed data are provided in Supplementary Table 2 and Supplementary Figure 1).

**Figure 1:**
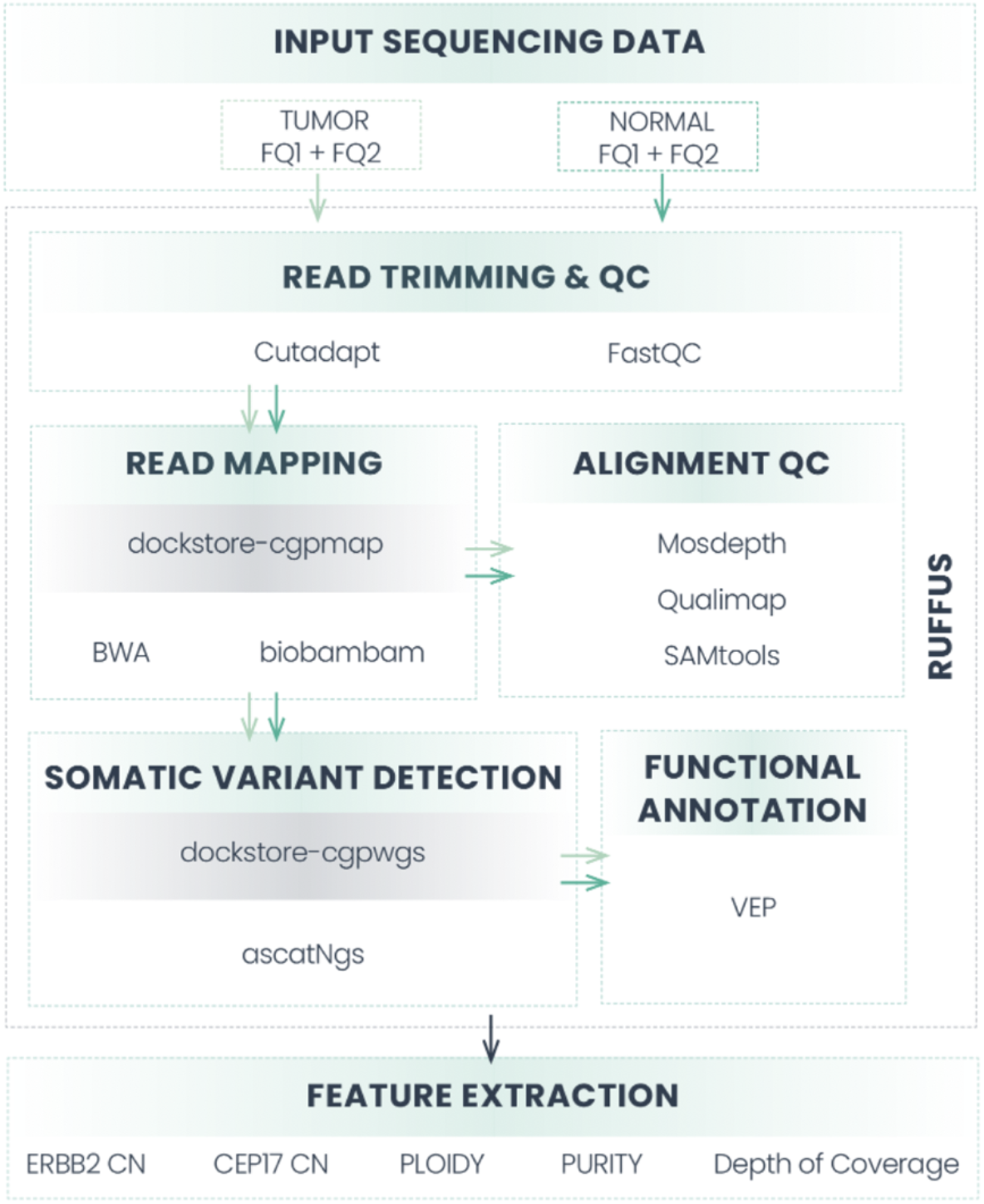
The summary of the in-house pipeline used for data extraction and processing.

Variant calling was performed using Sanger’s Cancerit CGPWGS pipeline v2.0.1 [20], and specifically copy number variants, purity, and ploidy were identified with ascatNgs [23]. Identified variants were annotated using Ensembl VEP v102 [24].

### 2.3 Analyzed parameters and method validation

In the study, we used only clinical data on HER2 status according to ASCO/CAP recommendations or, in the case of HMF metastatic/advanced tumours, the presence of targeted treatment with HER2 inhibitors, which was indicative of the confirmed presence of HER2 expression. Based on ASCAT copy number alteration calling, *ERBB2* (NC_000017.10:37844167_37886679) and uniquely mapped 8250 bp sequence adjacent to CEP17 (NC_000017.10:22236000_22244250) copy numbers were extracted along with ploidy and purity estimation for all the tumour samples. The data were used to create 3 features for HER2 status assessment: absolute ERBB2 CN, ERBB2_CN-n (ploidy-adjusted ERBB2 CN), and ERBB2_CN/CEP17_CN ratio. Based on these features, a machine learning-based classifier was constructed, which determined the best approach for HER2 status discrimination. 614 samples from the datasets were used as a training set, the remaining 264 samples served as a validation hold out set for the classifier and were not analyzed *a priori*.

A decision tree-based classifier was chosen after comparing the effectiveness of logistic regression, random forest and decision tree models. Decision tree outperformed other classifiers in terms of accuracy and interpretability.

For the decision-tree-based modelling, the discovery cohort was randomly split into a training (75%) and a test set (25%). The model was constructed on 3 aforementioned features and trained on the training set. Since the number of samples in IHC/FISH HER2-positive and negative groups was unbalanced (there were almost eight times less HER2+ samples than negative), we added class weights (8:1) to minimize the bias. After constructing the model we measured its performance on 264 samples from the validation set. We used accuracy, precision, and recall along with the F1-score. Cohen’s Kappa score was estimated to evaluate the non-randomness of classification.

To show how each of the 3 features influences the classifiers performance alone, we have established the same parameters independently for each of them as well and compared all the approaches with random data classification methods (Figure 2).

**Figure 2:**
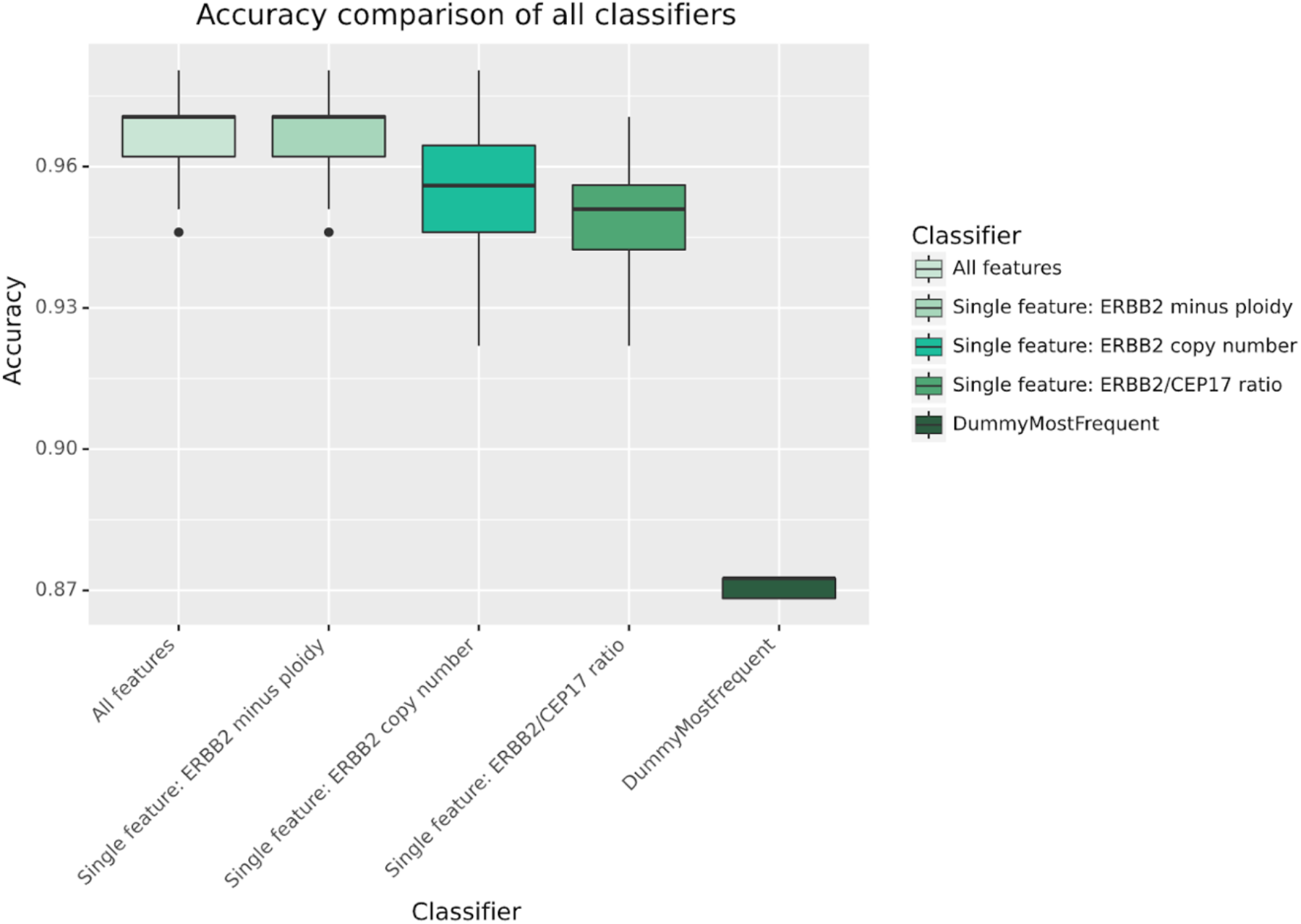
Accuracy comparison between 3 features used to determine HER2 amplification status in WGS data.

To further test the validity of our results, we decided to evaluate whether differences in tumour purity, heterogeneity of ploidy, or differences in mean depth of coverage had any deteriorative effects on the correctness of the results (see Supplementary Tables 2-3 and Supplementary Figures 1-2). For these experiments, we divided the samples into two near-equinumerous groups for each comparison and evaluated the differences in the tests’ performance. Finally, we determined the overall predictive value, PPV, and NPV with confidence intervals for the whole dataset of 877 genomes.

## 3. Results

In the analyzed dataset, 159 patients were categorized as triple-negative breast cancer (TNBC) (18%), among HER2-negative patients ER+/HER-accounted for 599 (88%). 110 (13%) of samples were identified by clinical testing as HER2-positive, among them: 74 ER+/HER2+ (8%), 36 ER-/HER2+ (4%). For 8 patients’ ER status was unavailable.

HER2 positivity was slightly underrepresented in favour of TNBC in comparison with statistics for the caucasian population (18%) which may be an accidental or sampling bias related to genomic consortia’s sample collection process, or an effect of discarding datasets with incomplete clinical data.

The decision tree machine learning approach has demonstrated the best discrimination between HER2-positive and negative cases based on a single parameter, ploidy corrected-ERBB2 CN with a threshold of 2.265 (fig.2). The decision tree algorithm was evaluated in 3-fold cross-validation repeated 10 times to estimate the mean value and standard deviation for each metric. The results were as follows: accuracy = 96,7% (+- 0.87%), precision = 86% (+- 5%), recall = 89% (+- 6%), Cohen’s Kappa = 85% (+- 3.7%) and F1 = 87% (+- 3%). A high value of Cohen’s Kappa strongly indicates that our model classifies samples in a non-random fashion.

The learning curve displayed no further improvement with sample numbers exceeding 150 instances, therefore we believe the results display the best reflection of the biological phenomenon of HER2 amplification we could extract from genomic data. Moreover, Principal Component Analysis of the dataset (fig.3) has shown a very good and robust separation of data into two groups, representing differences in HER2 status. As data distribution across depths of coverage, tumour purities, and ploidies were not normal, we decided to compare the accuracy distributions for these parameters with the Wilcoxon signed-rank test. The evaluation of results across data coverages has shown no significant differences (p>0.05) between groups.

**Figure 3:**
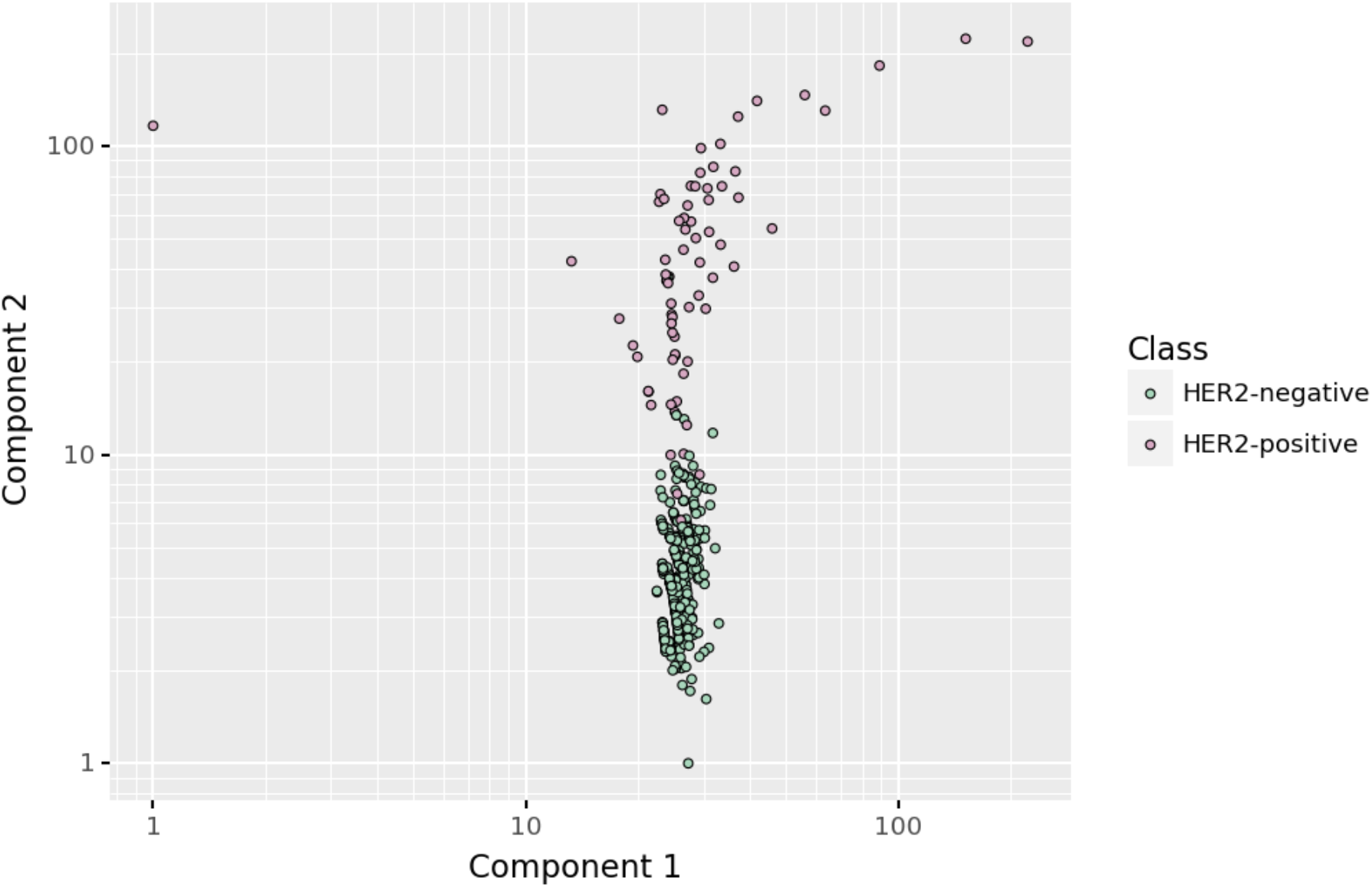
Principal Component Analysis of the dataset with 6 features: purity, Ploidy, ERBB2 CN, CEP17 CN, ERBB2 CN / CEP17 CN ratio, ploidy-corrected ERBB2 CN.

The comparison of low vs high purity also has not yielded significant differences (p>0.05). However, there is a significant decrease in mean accuracy of the test from 0.97 to 0.94, dependent on increased tumour ploidy above two (p=5.1×10^−6^) (Figure 4).

**Figure 4:**
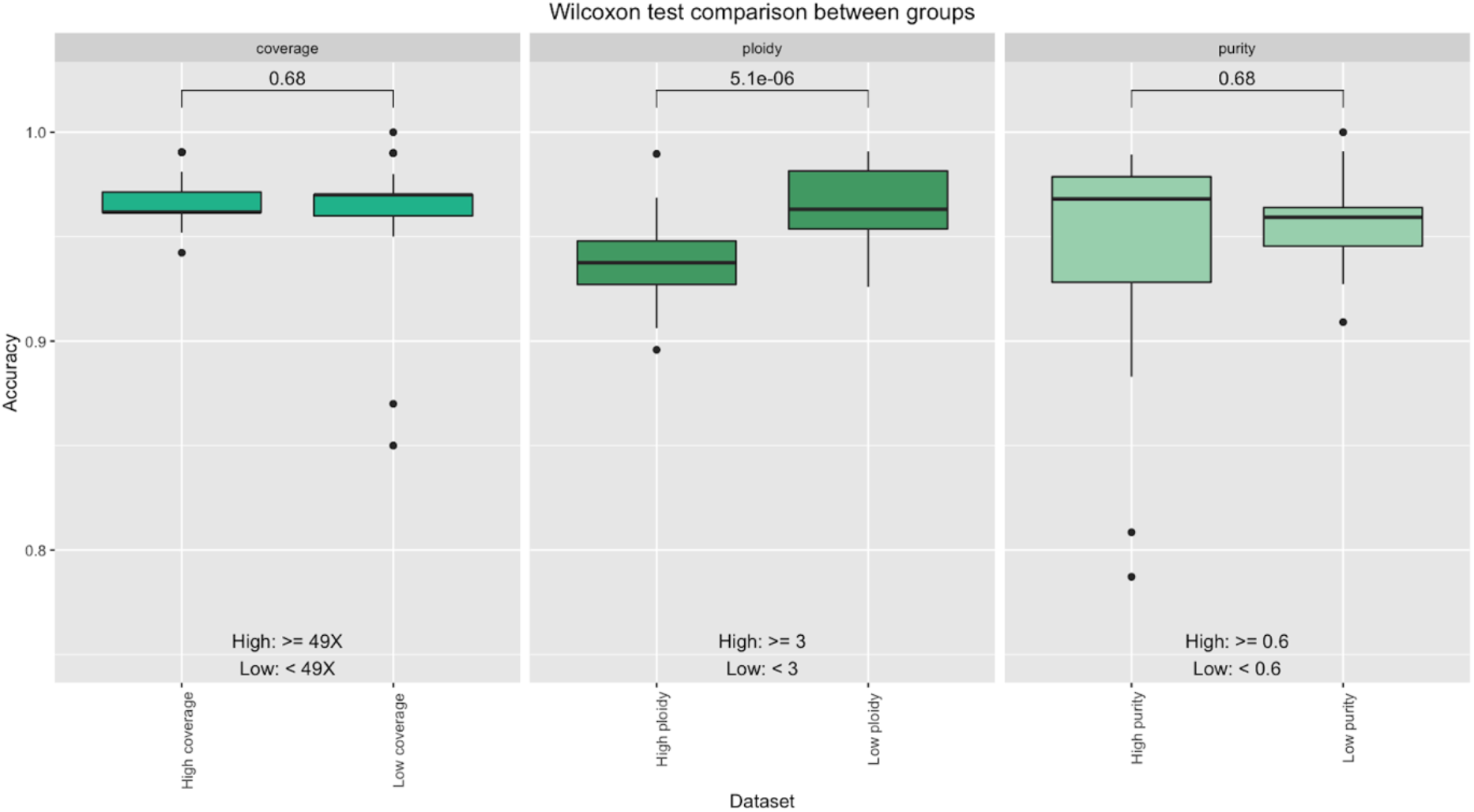
Wilcoxon test comparison of means between distributions of accuracies in: A) High vs low coverage data (threshold 49X), B) High vs low ploidy data (threshold 3), C) High and low purity data (threshold 0.6).

The analytical validation of the ploidy-corrected ERBB2_CN method gave the overall diagnostic sensitivity of 92.92% (95%CI 86.53-96.89%) and specificity of 97.91% (95%CI 96.62-98.8%) (Table 1).

**Table 1:** Analytical validation of the whole genome sequencing ploidy-corrected ERBB2 copy number. For details on samples used see Supplementary Data.

| IHC/FISH |  |  |
| --- | --- | --- |
| Non-Amplified | Amplified | WGS RESULTS |
| 748 | 8 | Non-Amplified |
| 16 | 105 | Amplified |

| Results | Point estimate | Lower CI | Upper CI |
| --- | --- | --- | --- |
| **Sensitivity** | 92.92% | 86.53% | 96.89% |
| **Specificity** | 97.91% | 96.62% | 98.80% |
| **Accuracy** | 97.16% | 95.83% | 98.15% |
| **Positive Predictive Value** | 88.67% | 82.78% | 92.73% |
| **Negative Predictive Value** | 98.74% | 97.57% | 99.35% |

## 4. Discussion

Decreasing next-generation sequencing prices and increasing availability of this technology in medical practice have encouraged the transition from conventional cytogenetic and molecular methods to NGS in oncology. However, the evidence on the reliability of NGS for clinical use in copy number detection is still very limited. As the HER2 protein is one of the most significant biomarkers for breast cancer diagnostics, prediction, and prognostics, there were several attempts to show the applicability of NGS techniques in this indication.

The largest analytical validation study was conducted by Memorial Sloan Kettering on their proprietary MSK-IMPACT Assay [8]. This hybrid-capture based panel NGS test was analyzed in 213 BC samples and evaluated in a clinical setting on further 599 BCs. The cutoff for positive result was established based solely on ERBB2 CN, adjusted to background and normal signal of diploid genomes (defined as ‘fold change’, FC=1.5). The group reported 95% specificity and 100% sensitivity on >10% of tumour content, with IHC/FISH evaluated by newest, 2018 guidelines and a dual-probe FISH assay [8]. Last year, a continuation of the study exploited the borderline cases with excellent concordance [12]. Several other studies have also proven the clinical value of panel NGS for HER2 testing in breast cancer and other solid tumours, using the same strategy of fold change determination, using either Illumina [1–4] or Ion Torrent methodology [5].

On the other hand, data on clinical whole genome sequencing utility is scarce. There have only been two small clinical validation studies with direct comparison to the orthogonal methods. The first, released by Hartwig Medical Foundation, was a part of a WGS pan-cancer validation study. The *ERBB2* status was evaluated on only 16 samples with overall concordance of 93%. HMF group compared ploidy and chromosome 17 CN with absolute CN of *ERBB2* but did not draw any conclusions due to the small sample size [5]. The second, performed by King’s College Hospital in London, was performed on 145 BC samples with only 27 positives for HER2. With the 4 samples discrepant, the sensitivity in the UK cohort was 88% and specificity 98% [25].

We attempted to systematically determine the criteria for whole genome sequencing of ERBB2 CN in matched-normal tumour samples. Our strategy was to gather publicly available breast cancer datasets with reliable clinical metadata and analyze them uniformly with minimal 20x depth of coverage. Our machine learning approach, based on Decision Tree classifier, objectively captured the superiority of ploidy-corrected ERBB2 CN over ERBB2 CN/CEP17 CN ratio and absolute ERBB2 CN for HER2 status evaluation in breast cancer by WGS. To measure the test’s reliability, we used Cohen’s kappa coefficient. The high value of 85% rules out the possibility of the data agreement occurring by chance.

This is, to the best of our knowledge, the first, the largest, and the most objective study of its kind, utilizing artificial intelligence and machine learning approaches for establishing diagnostic criteria. The optimal CAP/AAP guidelines for IHC/FISH testing took 11 years to refine, because it involved a series of consecutive evaluations and quality control rounds of diagnostic parameters which probably could have been done nowadays in an AI-based manner more robustly and quickly [6,7]. First proof-of-concept AI-based solutions for robust FISH and IHC assessment are already tested in clinical setup [26,27].

The AI/ML approach is an emerging field of medicine, improving the efficiency of pathomorphological assessment [28] radiology [29] and clinical chemistry [30]. In the field of breast cancer diagnostics, the genomics and transcriptomics is being applied to distinguish between intrinsic BC subtypes with different prognosis [31], identify new potential biomarkers or repurpose the existing. These strategies may only be used in the clinical setting after well-planned validation, showing concordance and stability of the test. Our results prove that WGS is a reliable method for HER2 clinical diagnostics, and it may be implemented as a standalone test or in combination with IHC instead of FISH or other NGS-based methods in routine practice. With diagnostic sensitivity of 92.92% and specificity of 97.91% determined on unselected and heterogeneous groups of patients, we conclude that the technology is mature and ready for prospective, multicenter analytical and clinical validation.

Our results do not deviate relevantly from those reported by other groups focused on HER2 NGS testing, however the diagnostic sensitivity is still not optimal. We suspect that heterogenous evaluation of IHC/FISH results, made on the basis of different issues of ASCO/AAP guidelines, may have contributed to the discrepancy. There was also great heterogeneity of the whole genome sequencing raw data, acquired on different equipment by different genomic consortia. There were also serious differences in tumour sample collection, DNA extraction, and library preparation methods. All these preanalytical and analytical factors must have contributed to the greater variation in HER2 results than in the single-facility method with uniform IHC/FISH evaluation methodology and a single laboratory protocol for sample management. Even so, the WGS method exhibits superb robustness and effectiveness, which is a great advantage, allowing for a low-cost external, even world-wide quality control assessment program to be held out in the near future.

Other factors contributing to slightly lower analytical sensitivity are changes in HER2 status, which could have occured in metastatic tumours from HMF dataset. In these instances, we couldn’t directly evaluate the correctness of IHC/FISH data, because they came from the primary biopsy, not the biopsy corresponding with the sample used for WGS. The shift between IHC positive and negative status is reported in up to 11.5% of HER2-negative cancers (conversion to HER2 positive) and in 37% of those initially positive (conversion to HER2 negative in presence of selective pressure of trastuzumab) [8,9].

Some of the discrepancies may have come from tumour subclonality, which is a common serious diagnostic issue. The signal from a small proportion of HER2 amplified cells may be below the resolution of whole genome sequencing at 30-60x depth of coverage [10].

The spatial intra-tumour heterogeneity may have also contributed to false negative/positive results when there were differences in sampling location between tissue collected for FFPE blocks and WGS (e.g., different distant metastases are sampled).

In addition, overexpression of HER2 is not always ERBB2-amplification based as about 5% of non-amplified tumours exhibit high overexpression. Even though there is currently no genomic background of this phenomenon known, whole genome sequencing could potentially detect alterations in HER2 regulatory pathways leading to overexpression, which could further improve WGS diagnostic power.

## 5. Conclusion

We provide evidence that the *ERBB2* status can be reliably determined by WGS methodology which may be included into a comprehensive test for breast cancer diagnostics. The 20% of tumour purity and 30x depth of coverage are sufficient to ensure good quality of genomic data in most instances. Given good concordance of a whole genome sequencing with routinely used methods, we suggest that assessment by a WGS method may be an alternative to other NGS-based methods as well as FISH-based diagnostic tools. Hence, it should be subjected for evaluation by ASCO/CAP in the future updates of the HER2 testing recommendations. In our work, we have also proven that short-reads WGS technology bears great potential for establishing a harmonized global quality assessment program for *ERBB2* detection, as the outputs of heterogeneous data gathered from 4 genomic consortia show a high degree of concordance between methodologies and pipelines.

## Data Availability

The data that support the findings of this study are openly available in the following repositories:
Hartwig Medical Foundation which was acquired under data request number DR-169,
International Cancer Genome Consortium, which was acquired under data request number DACO-6030,
The Cancer Genome Atlas data was acquired via dbGaP platform (project phs000178.v11.p8) under data request number #86794-3.
Secondary data that supports the findings of this study that was generated by the Authors are available in the supplementary material of this article.

https://www.hartwigmedicalfoundation.nl/

https://portal.gdc.cancer.gov/legacy-archive/search/f

## Abbreviations

WGS: whole genome sequencing,
HER2: human epidermal growth factor receptor 2
BC: breast cancer
CN: copy number
FISH: fluorescence in situ hybridization
IHC: immunohistochemistry
ASCO: American Society Cancer of Clinical Oncology
CAP: College American Pathologists
NGS: next-generation sequencing
CEP: Centromere enumeration probe
AI: artificial intelligence
TCGA: The Cancer Genome Atlas
ICGC: International Cancer Genome Consortium
HMF: Hartwig Medical Foundation
FFPE: formalin-fixed, paraffin-embedded
TNBC: Triple-negative breast cancer

## Data accessibility

The data that support the findings of this study are openly available in the following repositories:

Hartwig Medical Foundation at https://www.hartwigmedicalfoundation.nl/ which was acquired under data request number DR-169.

International Cancer Genome Consortium at https://dcc.icgc.org/pcawg/which was acquired under data request number DACO-6030.

The Cancer Genome Atlas data was acquired via dbGaP platform (project phs000178.v11.p8) at https://www.ncbi.nlm.nih.gov/projects/gap/cgi-bin/study.cgi?study_id=phs000178.v11.p8, under data request number #86794-3. Secondary data that supports the findings of this study that was generated by the Authors are available in the supplementary material of this article.

## Acknowledgements

We thank the international genomic consortia, their researchers and public funding institutions who have generously donated the genomic data and clinical metadata for this project.

This publication and the underlying study have been made possible by the data that the Hartwig Medical Foundation and the Center of Personalized Cancer Treatment (CPCT) have made available. The results published here are also in part based upon data generated by the TCGA Research Network: https://www.cancer.gov/tcga.

This work was also supported by data obtained from ICGC Breast Cancer Working Group and The Cancer Genome Atlas.

Importantly, we thank the patients and their families for their participation in the genomic projects and the opportunity to use their clinical and genomic data for fundamental cancer research. We would also like to acknowledge the work of all clinical staff gathering samples and medical records. Lastly, we thank all members of Adam Mickiewicz University, Poznan Supercomputing and Network Centre and MNM Diagnostics staff for help with genomic data processing and general support.

## Contribution

Wojtaszewska M-first author, study design, manuscript preparation, data analysis. Wozna A-genomic data administration, study design.

Piernik M -pipeline development, bioinformatics support, design of the computational framework for machine learning approach.

Dabrowski M-data refinement, supporting in writing and corrections of the manuscript.

Gniot M-statistics, genomic data administration, manuscript corrections.

Szymański S - curation of medical data.

Socha M – statistics, pipeline refinement, data analysis

Kasprzak P^-^ expertise on molecular HER2 status assessment, clinical data evaluation.

Matkowski R-expertise on molecular HER2 status assessment, clinical data evaluation.

Zawadzki P^1,3^ - supervision of the project, review of methodology and manuscript.

## Supporting information

### Supplementary Information

Detailed methodologies of Whole Genome Sequencing used by main Genomic Consortia and input data heterogeneity.

Supplementary Table 1: Differences in DNA preparation, library preparation and sequencing technologies across Genomic Consortia.

Supplementary Table 2: Coverage heterogeneity across datasets

Supplementary Table 3: Purity and ploidy heterogeneity across datasets

Supplementary Figure 1: Tumour (A) and Normal (B) coverage heterogeneity across Consortia.

Supplementary Figure 2: Purity (A) and ploidy (B) heterogeneity across Consortia.

### Supplementary Data

List of samples used in the study classified by IHC/FISH and WGS and additional list of discarded samples.

