## Supplementary Information for "Validation of HER2 status in whole genome sequencing data of breast cancers with AI-driven, ploidy-corrected approach"

**Supplementary information: Detailed methodologies of Whole Genome Sequencing used by main Genomic Consortia and data heterogeneity.**

### A. WGS methodology details

| | Material | ASCO/CAP HER2 recommendations issue | DNA extraction | DNA fragmentation | Insert size | Library preparation | Read mode | Sequencing platform |
|---|---|---|---|---|---|---|---|---|
| HMF [1] | Biopsy (core needle from metastasis, or LN frozen primary tumor) | 2013/2018 | DSP DNA Midi, Qiagen | Sonication (Covaris) | 450 | TruSeq Nano LT | 2x150 | HiSeqX/ NovaSeq6000 |
| | blood | - | Qiasymphony DSP DNA mini, Qiagen | | | | | |
| TCGA [2] | Frozen primary tumor | 2007 | DNA/RNA Allprep, Qiagen | Bead Linked Transposomes | 300-350 | Illumina Nextera DNA Flex Library Preparation Kit | 2x150 | NovaSeq6000 |
| | blood | - | DNA Midi, Qiagen | | | | | |
| ICGC [3] | Breast cancer tissue | 2013 | NA | Sonication (Covaris) | 500 | TruSeq Nano LT | 2x100 | HiSeq 2000/ HiSeq 2500 |
| | Blood lymphocytes, skin or breast tissue | - | NA | | | | | |

Supplementary Table 1: Differences in DNA preparation, library preparation and sequencing technologies across Genomic Consortia.

Data source:


1) Priestley, P., Baber, J., Lolkema, M.P. *et al.* Pan-cancer whole-genome analyses of metastatic solid tumours. *Nature* **575,** 210–216 (2019). https://doi.org/10.1038/s41586-019-1689-y
2) Rossing, M., Sørensen, C. S., Ejlertsen, B., & Nielsen, F. C. (2019). Whole genome sequencing of breast cancer. *APMIS : acta pathologica, microbiologica, et immunologica Scandinavica*, *127*(5), 303–315. https://doi.org/10.1111/apm.12920
3) Nik-Zainal, S., Davies, H., Staaf, J. *et al.* Landscape of somatic mutations in 560 breast cancer whole-genome sequences. *Nature* **534,** 47–54 (2016). https://doi.org/10.1038/nature17676

### B. Coverage heterogeneity of input data

| | Average tumour sample coverage (min-max; SD) | Average normal coverage (min-max, SD) |
|---|---|---|
| **All datasets** | 48,27 (27,20-75,50; 4,.31) | 36,03 (25,12-63,47; 5,37) |
| **HMF (downsampled)** | 49,08 (40,40-52,67; 1,85) | 36,59 (25,12-59,21; 5,25) |
| **ICGC** | 43,23 (28;23-75,5; 7,69) | 35,58 (26,24-63,47; 5,85) |
| **TCGA** | 48,96 (27,20-59,00; 7,05) | 31,24 (26,46-39,95; 2,63) |

Supplementary Table 2: Coverage heterogeneity across datasets

**A** **B**

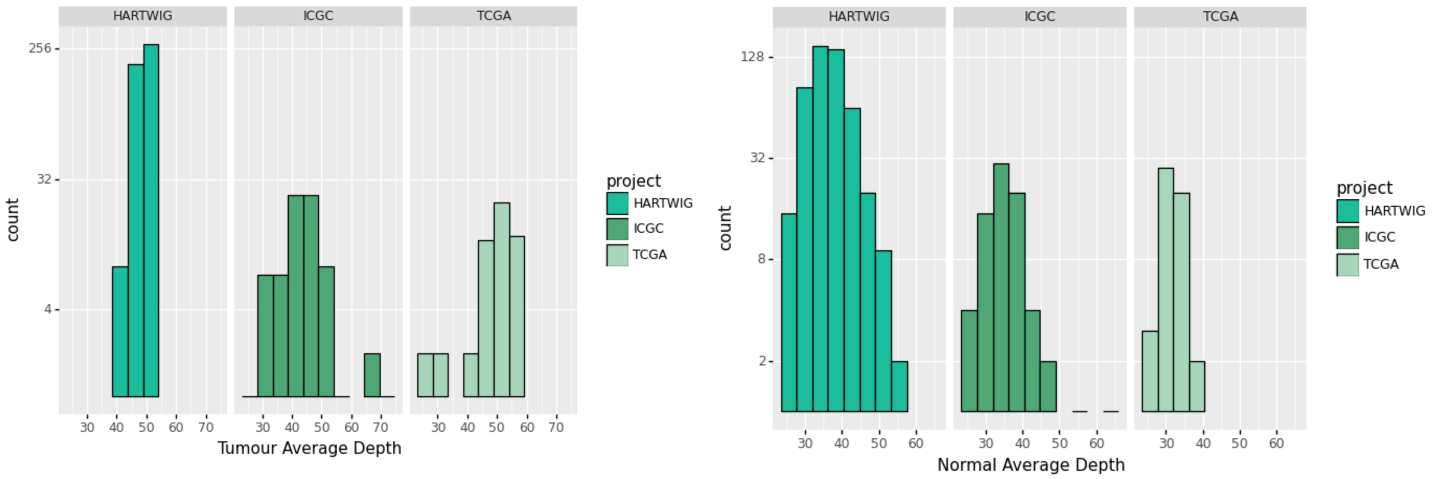


Supplementary Figure 1: Tumour (A) and Normal (B) coverage heterogeneity across Datasets. <u>HMF data were downsampled by the Authors.</u>

### C. Heterogeneity in tumour purity and ploidy across Consortia

| | Average tumour sample purity (min-max; SD) | Average tumour ploidy (min-max, SD) |
|---|---|---|
| **All Consortia** | 0,56 (0,2-1; 0,18) | 36,03 (1,51-5,54; 5,37) |
| **HMF** | 0,56 (0,2-1; 0,19) | 2,84 (1,51-5,54; 0,86) |
| **ICGC** | 0,53 (0,21-1; 0,19) | 2,74(; 0,86) |
| **TCGA** | 0,58 (0,21-0,92) | 3,24 (1,79- 5,54; 0,84) |

Supplementary Table 3: Purity and ploidy heterogeneity across datasets

**A** **B**

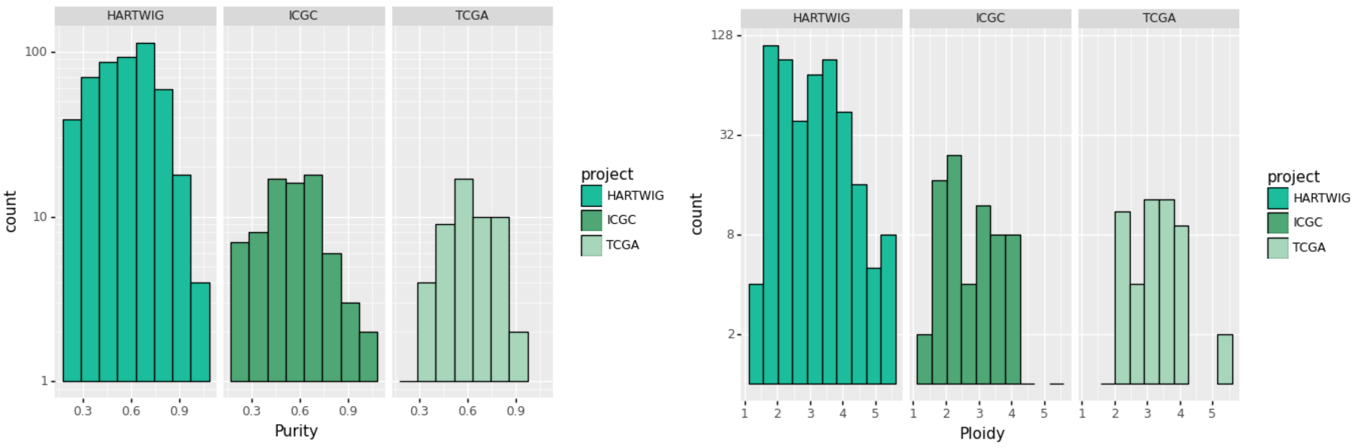


Supplementary Figure 2: Purity (A) and ploidy (B) heterogeneity across Consortia.
